# Diffusion MRI Profiles Map onto Distinct Inflammatory States After Adolescent Concussion: A CARE4Kids Study

**DOI:** 10.64898/2026.07.17.26358354

**Authors:** Arum Lim, Jessica M Gill, Kevin C Bickart, Adrian I Onicas, Jeffrey J Bazarian, John Alice, Christine L Mac Donald, Anne Brown, Lawrence Cook, Frederick P Rivara, Gerald A Gioia, Christopher C Giza, Emily L Dennis, the Concussion Assessment, Research, and Education for Kids (CARE4Kids) Consortium

## Abstract

**Importance:** Neuroinflammation is a key component of the response to injury after concussion, but direct links between diffusion MRI metrics and specific plasma inflammatory pathways in human concussion have not been established.

**Objective:** To examine associations between diffusion MRI metrics and pathway-level inflammatory proteomic signatures in adolescents during the subacute period after concussion.

**Design, Setting, and Participants:** Cross-sectional analysis of data from the CARE4Kids Consortium, a six-site prospective study. Participants were English-speaking adolescents ages 11–17.99 with concussion and symptoms at 7–35 days post-injury. Data were collected between 2022-2024. Of 370 enrolled participants, 122 had both diffusion MRI and plasma proteomics available for analysis.

**Exposure:** Advanced diffusion MRI metrics were converted to z-scores and participants were grouped by the spatial extent of outlier values (potholes and peaks) across 15 white matter regions of interest. Nine non-redundant groupings were selected for primary analysis.

**Main Outcomes and Measures:** Pathway-level inflammatory profiles derived from gene set enrichment analysis (GSEA) of ∼5,400 plasma proteins measured by Olink proximity extension assay, targeting nine hallmark inflammatory pathways spanning initiation through resolution. Persistent symptoms were assessed 64-115 days post-injury.

**Results:** Diffusion metrics reflecting tissue disorganization were associated with upregulation of the coagulation pathway, consistent with hemostatic-inflammatory signaling. Metrics reflecting reduced tissue complexity and neurite density were associated with upregulation of interferon-α and interferon-γ response pathways, consistent with microstructural remodeling driven by cellular immune activation. Elevated free water content was associated with downregulation of most inflammatory pathways and trend-level transforming growth factor - β upregulation, reflecting inflammatory resolution. Time since injury did not differ between groups based on free water (Kolmogorov-Smirnov p = 0.97), suggesting these differences reflect individual variability in recovery pace. Exploratory analyses showed a trend toward lower odds of persistent symptoms in the group with elevated free water content (odds ratio = 0.51, *p* = 0.18).

**Conclusions and Relevance:** Multiple diffusion MRI metrics are differentially sensitive to distinct neuroinflammatory states in the subacute period after adolescent concussion. These findings suggest that diffusion imaging could serve as a non-invasive tool for inflammatory phenotyping, with potential implications for identifying patients who may benefit from targeted immunomodulatory intervention.

**Key Points:** *Question:* Are specific diffusion MRI metrics sensitive to distinct inflammatory pathway signatures in adolescents during the subacute period after concussion?

*Findings:* In this cross-sectional study of 122 adolescents with concussion, multimodal diffusion MRI metrics were associated with mechanistically distinct proteomic signatures rather than a uniform inflammatory response. Metrics reflecting tissue disorganization were associated with hemostatic-inflammatory pathway upregulation, metrics reflecting reduced tissue complexity and neurite density were associated with interferon signaling and coagulation suppression, and elevated free water content was associated with inflammatory pathway downregulation and trend-level resolution-phase signaling.

*Meaning:* Different diffusion MRI metrics are sensitive to qualitatively different neuroinflammatory processes, suggesting that multimodal diffusion imaging may enable non-invasive profiling of the post-injury inflammatory state and inform targeted therapeutic strategies.

## Introduction

Concussions affect ∼1.9 million U.S. adolescents annually, with 15-30% developing persistent post-concussive symptoms (PPCS) lasting more than 3 months.^1,2^ Current demographic and symptom-based prediction methods have limited ability to identify biological contributors to prolonged recovery.^3,4^ Multiple mechanisms have been proposed — diffuse axonal injury, neurometabolic dysfunction, and biopsychosocial factors^5,6^ — but prolonged neuroinflammation has emerged as a key contributor. Inflammation after concussion is initiated by primary injury and activates microglia and other immune cells to clear damage and facilitate repair,^7^ but over-activation can clear undamaged neurons and produce white matter (WM) loss.^8,9^ Immune activation also initiates neuropeptide signaling to support neuronal repair and restore function.^10,11^ Clinical studies confirm cytokine elevation after TBI,^12,13^ generally tracking severity and partially predicting recovery.

Neuroinflammation is not readily detectable on standard or advanced neuroimaging. Diffusion magnetic resonance imaging (dMRI) has emerged as one of the most sensitive markers of disruption after TBI.^14^ dMRI measures the diffusion of water in the brain, making it sensitive to processes that disrupt the organized movement of water along WM fibers, including axonal damage, demyelination, and inflammation.

Our understanding of when inflammation crosses from beneficial to detrimental is incomplete,^15^ in part because direct measurement is difficult. Lumbar puncture and translocator protein positron emission tomography (TSPO PET) are rarely performed even in severe TBI, both are impractical and ethically problematic in healthy children. Recent work suggests TSPO PET may indicate microglial density in humans rather than activation.^16^ Peripheral blood markers offer an alternative window into the neuroinflammatory response, though they reflect systemic rather than brain-specific processes. Standard diffusion tensor imaging (DTI) yields metrics like fractional anisotropy (FA), with decreased FA linked to IL-1β elevation in depression.^17^ Diffusion kurtosis imaging (DKI) extends DTI by capturing non-Gaussian diffusion, with kurtosis metrics associated with reactive astrogliosis,^18^ Crohn’s inflammatory activity,^19^ and both histological and blood markers of inflammation in animal models.^20^ Neurite Orientation Dispersion and Density Imaging (NODDI)^21^ explicitly separates intracellular, extracellular, and free water compartments, yielding intracellular volume fraction (ICVF; neurite density), orientation dispersion index (ODI; fiber coherence), and isotropic volume fraction (ISOVF; free water/edema). Elevated ISOVF early after TBI has been interpreted as vasogenic edema,^22^ ODI has been associated with C-reactive protein in healthy aging,^23^ and IL-6 expression in mice produces NODDI-detectable WM changes.^24^ These findings suggest different diffusion models may be sensitive to different facets of the neuroinflammatory response.

Few studies have examined diffusion-inflammation relationships in adolescents, where developmental variability complicates interpretation, and most existing work focuses on the acute or chronic phase rather than the subacute period when patients seek clinical care. A recent study combining MRI with proteomic markers identified distinct biomarker clusters that differed in WMmicrostructure, with higher FA and elevated C-C motif chemokine ligand 2 (CCL2) characterizing an acute inflammatory phenotype.^25^ In remote TBI, higher inflammatory cytokines have been linked to lower FA and greater symptom burden, suggesting chronic inflammation may contribute to lasting WM changes.^26^ Clinical trials using broad anti-inflammatory agents following TBI have failed to demonstrate benefit,^27^ underscoring the need to map imaging metrics onto specific inflammatory phenotypes.

The present study addresses these gaps by combining multimodal diffusion MRI with pathway-level proteomic profiling using a high-throughput proteogenomic sequencing method in adolescents during the subacute period after concussion. We leverage data collected by the CARE4Kids Consortium to comprehensively investigate whether dMRI metrics are associated with inflammatory pathways. We hypothesize that dMRI metrics will be associated with inflammatory pathways, and that different metrics will be sensitive to different components of the inflammatory response.

## Methods

### Participants

Participants were enrolled at each of the six sites in the C4K Consortium (https://www.care4kidsstudy.com/). Sites for recruitment included primary care clinics, emergency departments, concussion subspecialty clinics, athletic leagues, and schools. Eligible participants included English speakers ages 11-17.99 years diagnosed with a concussion by a health care provider using the Concussion in Sport Group criteria. Additionally, participants were included only if they continued to experience post-concussion symptoms at T1 (7-35 days after injury). Participants with a history of developmental disorders, neurological disorders, severe psychiatric illness, substance abuse, moderate or severe TBI, or prior concussion within the last 3 months were excluded from the study. Protocol was approved by the University of Utah institutional review board (IRB) as single IRB; participants assented and parents provided written informed consent. Further study details have been previously published.^28^

### MRI Acquisition

dMRI was collected at five of the six sites, with acquisition parameters in **Supplementary Table 1**.

### Blood Sample Acquisition and Preparation

Non-fasting peripheral blood samples were collected at T1 (study enrollment). Plasma was collected from venous whole blood within 30 min of blood collection by centrifugation at 2000 × g for 15 min, stored locally at −80°C, and shipped overnight on dry ice to the biomarker repository for long-term storage.

### MRI Processing

Diffusion-weighted images were preprocessed using a standard pipeline including MP-PCA denoising, Gibbs ringing removal, susceptibility distortion correction, and eddy current and motion correction (FSL [FMRIB Software Library],^29^ MRtrix3,^30^ DIPY [Diffusion Imaging in PYthon],^31^ dmipy [diffusion microstructure imaging in python]^32^). DTI, DKI, and NODDI models were fit to derive 12 metrics (FA mean diffusivity [MD], axial diffusivity [AD], radial diffusivity [RD], kurtosis FA [KFA], mean kurtosis [MK], axial kurtosis [AK], radial kurtosis [RK], ICVF, ODI, and ISOVF). Full preprocessing details are provided in **Supplementary Methods.**

### Neuroimaging Groupings

Processed ROI values were harmonized across sites using ComBat^33^ and adjusted for age, sex, and time since injury, then converted to z-scores based on the study sample (**Figure 1**). For each participant and diffusion metric, we counted the number of ROIs with z-scores below −1 (“potholes”) or above +1 (“peaks”). Participants were assigned to one of three ordinal groups for each metric, separately for potholes and peaks: Group 0 (no extreme-valued ROIs), Group 1 (at or below the 75th percentile of the nonzero distribution), or Group 2 (above the 75th percentile, reflecting widespread atypical values). A histogram showing the distribution of potholes and peaks for each measure with the corresponding 75th value is shown in **Supplementary Figure 1**. This was applied across 12 imaging metrics, producing 24 groupings. Pairwise correlations (**Supplementary Figure 2**) were used to select 9 non-redundant groupings for primary analysis: AD_low, AK_low, FA_low, ICVF_low, ISOVF_high, ODI_high, RD_high, RK_low, and RDI_high. **Supplementary Figure 3** displays how frequently each ROI appears as a pothole or peak.

**Figure 1.**
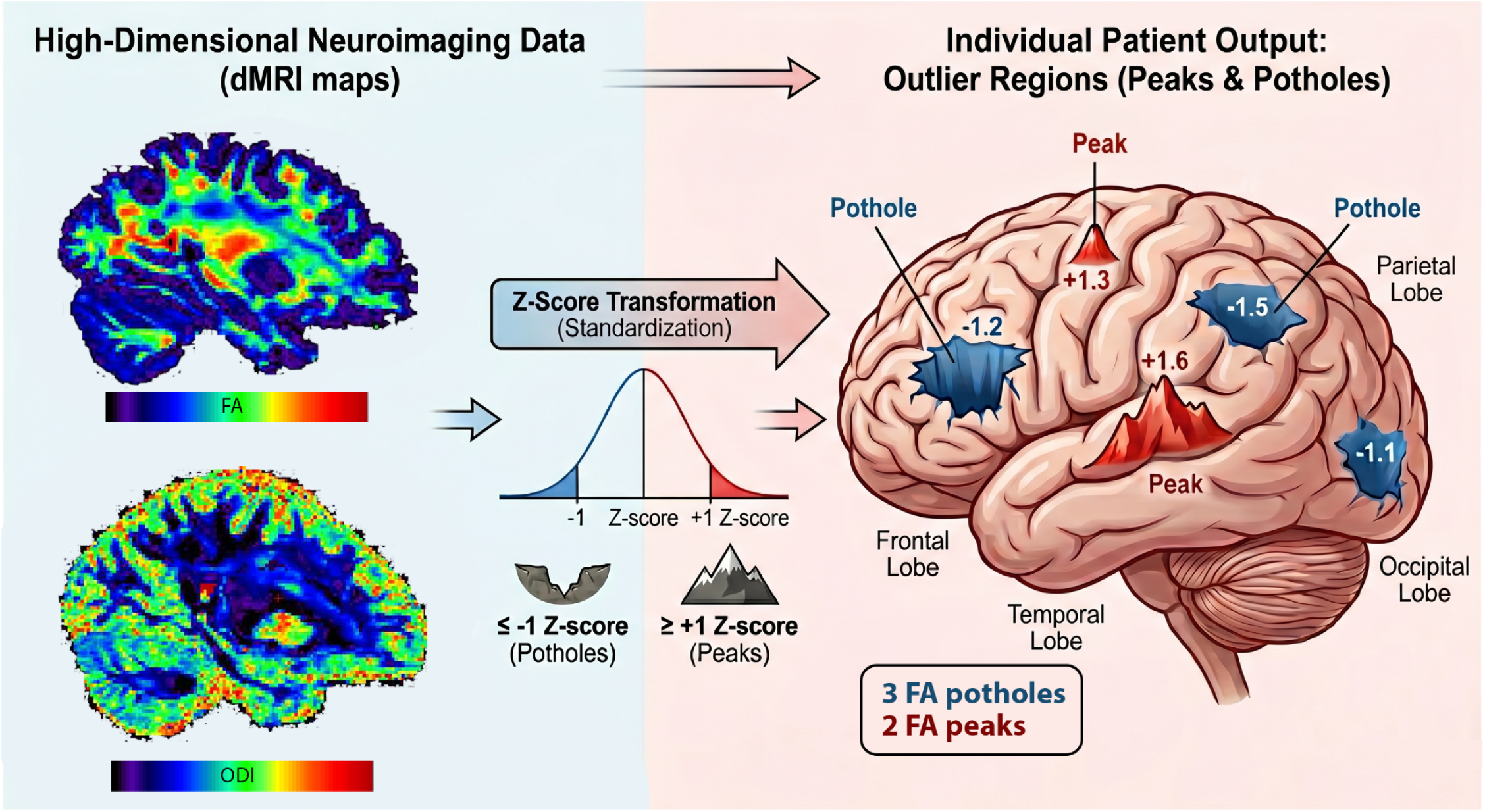
Potholes and peak imaging analysis. Schematic demonstrating the process through which high dimensional dMRI data were converted to simple counts of potholes and peaks as described in the methods. Voxelwise maps of fractional anisotropy (FA) and orientation dispersion index (ODI) are shown on the left, with a schematic representing the individual output on the right.

### Proteomic Data Processing

Olink Explore HT was used to assess the relative concentration of approximately 5400 proteins for exploratory purposes. Olink is a high-throughput proteomics platform using Proximity Extension Assay (PEA) technology to enable precise, multiplex protein biomarker analysis. Employing dual antibody binding and DNA-based detection, it offers high specificity and sensitivity in biomarker detection. Details on Olink processing may be found in **Supplementary Methods.**

### Differential Protein Expression Analysis

Differential protein expression was assessed between the MRI-derived group classifications using a non-parametric Wilcoxon rank-sum test. Multiple comparisons were adjusted using the Benjamini-Hochberg false discovery rate, yielding q-values (adjusted *p*-values).

### Gene Set Enrichment Analysis

Proteins were ranked using signed -log10(*p*-value) to capture both the direction of fold changes and statistical significance from the differential analysis. Pre-ranked Gene Set Enrichment Analysis (GSEA) was performed using the *fgsea* package in R (ver 4.4.3) for *a priori* selected nine inflammatory gene sets from the Hallmark pathways: Tumor necrosis factor (TNF)-α signaling via Nuclear factor kappa B (NFKB), Interleukin (IL) 6_Janus Kinases (JAK)_Signal Transducers and Activators of Transcription (STAT) 3 signaling, IL2_STAT5 signaling, inflammation response, Complement, Coagulation, Interferon (IFN)-α response, IFN-γ response, and Transforming Growth Factor (TGF)-β signaling. Pathways with significant enrichment were identified using normalized enrichment scores (NES) and q-values (<.25, <.05, <.01, or <.001). The findings were summarized and presented as a heatmap, which allows comparison of NES for gene sets across the nine MRI metrics.

### Ingenuity Pathway Analysis

Protein networks for a subset of dMRI metrics with distinct pathway profiles were generated using Ingenuity Pathway Analysis (IPA, QIAGEN). Network visualization conventions are described in **Supplementary Methods**.

### Persistent Postconcussive Symptoms

Symptoms were assessed approximately three months after injury (range 64-115 days) using the PCSI-2, with persistent postconcussive symptoms (PPCS) defined as having a PCSI-2 Total Rapid Score (self-reported pre-injury PCSI-2 symptom score subtracted from the self-reported post-injury PCSI-2 score) greater than 6 at the 3-month visit.^34^

## Results

### Participants

370 adolescents were recruited (M age = 15.0, SD = 1.8 years; 54% female), with 200 completing MRI scans. Six participants were scanned but dMRI was not collected. Of 194 scans, 11 had acquisition errors and 1 was removed for motion-related quality issues, leaving 182 usable scans. Of the 182 participants with dMRI data, 122 also had blood samples collected at T1. Demographic and clinical variables for the subsets (neuroimaging, neuroimaging+blood) are shown in **Table 1**. None of the demographic or clinical variables differed significantly between subsets. Consort diagram shown in **Figure 2**.

**Figure 2.**
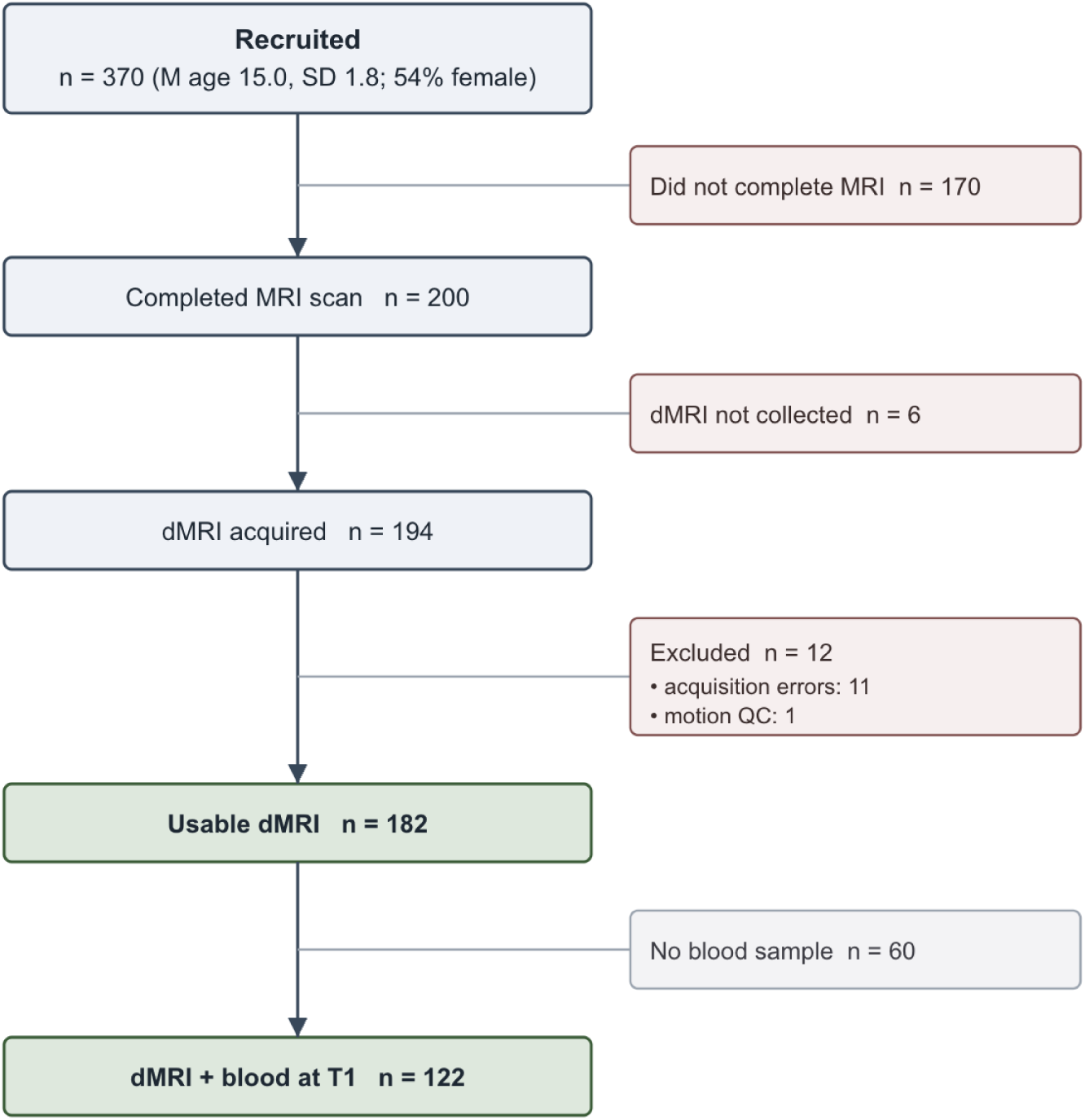
Flow of participants from recruitment to the analytic samples. Retained samples are shown in the main column; exclusions at right.

**Table 1.** Sample characteristics. The sample size, sex distribution (male | female), age in years (mean and standard deviation [SD]), time since injury in days (TSI, mean and SD), % of sample with persistent post-concussive symptoms (PPCS) at 3 months post-injury, race (%), ethnicity (%), and injury mechanisms (%) for the full sample, sample with dMRI data, and sample with both dMRI and Olink data are listed. No significant differences were observed between the dMRI or dMRI+Olink subsets and the full sample on any characteristic (all p > 0.05).

|  | Full Sample<br>(N=370) | dMRI Subset<br>(N=182) | dMRI + Olink Subset<br>(N=122) |
| --- | --- | --- | --- |
| Sex (M \| F) | 168 \| 202 | 92 \| 90 | 66 \| 56 |
| Age, mean (SD) | 15.0 (1.8) | 14.9 (1.9) | 14.7 (1.9) |
| TSI, mean (SD) | 21.2 (8.5) | 21.4 (8.4) | 21.4 (8.4) |
| PPCS at 3 months | 41.6% | 40.1% | 44.3% |
| **Race** |  |  |  |
| Asian | 3.8% | 2.7% | 2.5% |
| Black | 14.0% | 9.9% | 9.8% |
| White | 62.2% | 65.4% | 64.7% |
| More than one or other | 11.1% | 11.0% | 11.5% |
| Unknown or not reported | 8.9% | 11.0% | 11.5% |
| **Ethnicity** |  |  |  |
| Latino | 26.8% | 28.4% | 29.5% |
| **Injury Mechanism** |  |  |  |
| Sport/recreation | 63.2% | 62.1% | 59.0% |
| Fall | 10.5% | 11.5% | 14.8% |
| Struck by/against object | 10.5% | 12.1% | 10.6% |
| Motor vehicle collision | 9.5% | 8.8% | 9.0% |
| Assault | 3.8% | 3.3% | 3.3% |
| Other/missing | 2.4% | 2.2% | 3.3% |

### Olink Analysis

The heatmap summarizes NES across MRI-defined groups, providing an overview of pathway-level differences (**Figure 3**). Corresponding enrichment plots illustrate how proteins are distributed within the ranked list for selected significant pathways, demonstrating the degree and direction of coordinated enrichment (**Supplementary Figure 4**).

**Figure 3.**
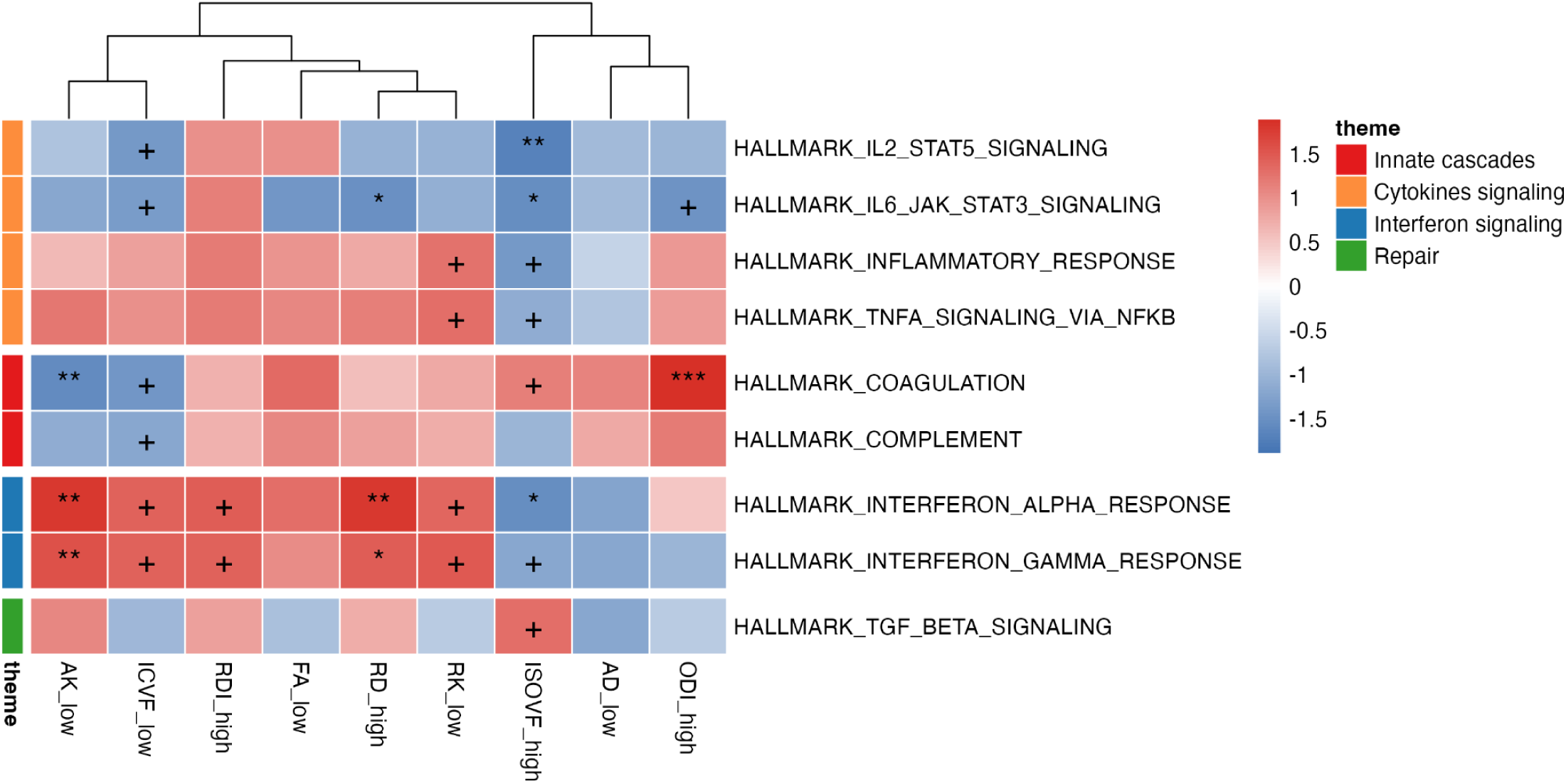
Heatmap of normalized enrichment scores. Colors represent normalized enrichment scores (NES). Positive values (red) indicate enrichment in cases and negative values (blue) indicate enrichment in comparisons. Asterisks denote pathways meeting the predefined FDR threshold (+<0.25 *<0.05, **<0.01, ***<0.001).

Proteins involved in coagulation were collectively shifted toward higher expression in the ODI_high group, suggesting coordinated activation of coagulation-related inflammatory processes in participants with elevated ODI. The ISOVF_high group showed enrichment for proteins involved in the coagulation and TGF-β signaling pathways. In contrast, the AK_low and ICVF_low groups exhibited higher expression of proteins involved in IFN-α and IFN-γ response pathways, alongside relatively lower expression of complement and coagulation-related proteins.

Network analysis in the AK_low group shows a protein cluster associated with cardiovascular system development and function, lymphoid tissue structure and development, and tissue morphology (**Figure 4a**). This protein network implicates phosphoinositide 3-kinase (PI3K)/JAK-mediated inflammatory dysregulation and GFAP upregulation, indicating astrocytic injury in the context of the low-AK metric. A protein network in the ISOVF_high group was characterized by humoral immune responses, inflammatory responses, and organismal injury and abnormalities (**Figure 4b**). Under high ISOVF, downregulation of immune surveillance machinery (IL15RA, CD83, CD55) and upregulation of NOTCH2, involved in angiogenesis, were observed.

**Figure 4.**
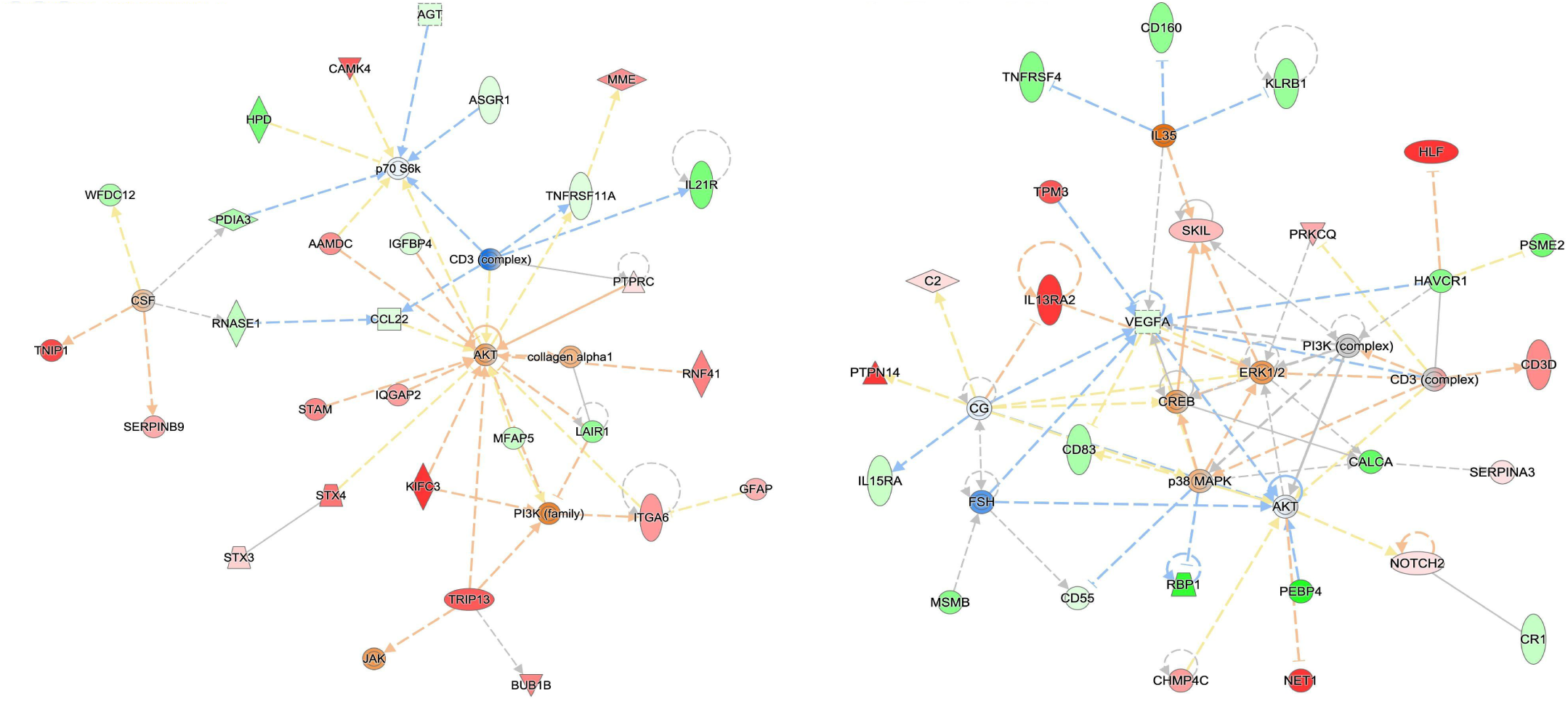
Networks of differentially expressed proteins in the AK_low group (A) and in the ISOVF_high group (B). (A): Cardiovascular System Development and Function; Lymphoid Tissue Structure and Development; Tissue Morphology. (-log10p=46); (B): Humoral Immune Response; Inflammatory Response; Organismal Injury and Abnormalities. (-log10p=49)

In an exploratory analysis, participants in the ISOVF_high Group 2 showed a trend towards a lower rate of PPCS than Group 0 (33% vs. 43%; OR = 0.51 [95% CI:0.19,1.37], *p* = 0.18 after adjusting for age, sex, and TSI), though this did not reach statistical significance.

## Discussion

Here we present a comprehensive analysis linking dMRI metrics to inflammatory pathways. A central finding is that diffusion MRI metrics do not map uniformly onto a single inflammatory response; instead, individual metrics are associated with mechanistically distinct inflammatory states.

Interferon signaling pathways demonstrated the broadest correspondence with dMRI metrics, with upregulation of both IFN-α and IFN-γ across groupings based on AK, RD (q<.05), RK, ICVF, and RDI (q<.25). The constellation of metrics suggests active microstructural remodeling: low AK and RK indicate reduced tissue complexity, low ICVF reflects decreased neurite density, and high RD suggests greater perpendicular diffusion, collectively consistent with clearance of myelin debris and cellular remnants. High RDI may reflect a simplified axonal compartment with persisting extracellular crowding from inflammatory infiltrates, producing a dissociation between reduced kurtosis and increased restriction.

AK_low exhibited a strong interferon profile, with significant enrichment of IFN-α and IFN-γ response pathways alongside relative suppression of coagulation proteins. IFN-α and IFN-γ activate microglia from a surveillant to a phagocytic (M1-like) phenotype that peaks a week after TBI and persists for several weeks.^15,35^ IFN-γ is also a well-established driver of oligodendrocyte injury and myelin thinning,^36^ thereby reducing axonal compartmental complexity and lowering kurtosis. Protein networks for AK_low were anchored by an AKT/JAK hub with GFAP elevation and CD3 engagement, consistent with sustained glial injury and innate-to-adaptive immune crosstalk. Co-occurrence of AK_low with ICVF_low further supports this interpretation, as interferon-driven activation would simultaneously clear axonal contents and decrease neurite density as part of the same remodeling process. Relative suppression of coagulation and complement was also observed, potentially mediated by regulatory feedback whereby IFNs inhibit these pathways.^37^ Longitudinal studies are needed to determine whether this interferon-dominant profile represents a distinct injury subtype or a later stage in a common inflammatory cascade.

The ODI_high group was characterized by upregulation of the coagulation pathway. High ODI reflects loss of coherent WM organization that can result from edema, inflammatory infiltration, or gliosis. When pathophysiological states disrupt the blood-brain barrier, complement activation initiates coagulation cascades through tissue factor exposure,^38^ and the resulting complement and coagulation factors can modulate neuronal conduction and synaptic transmission.^38,39^ The association between ODI_high and coagulation upregulation suggests that participants with widespread loss of fiber coherence show evidence of hemostatic activation, potentially reflecting vascular compromise and extravasation of coagulation proteins into the brain parenchyma.

The ISOVF_high group exhibited a distinct proteomic profile characterized by downregulation of several inflammatory pathways alongside trend-level upregulation of TGF-β signaling. High ISOVF reflects elevated free water content in WM, which can arise from expanded extracellular space as inflammatory cells clear and edema resolves. The simultaneous downregulation of cytokine and interferon signaling suggests coordinated immunoregulation consistent with TGF-β, which suppresses JAK/STAT, NF-kB, and interferon signaling.^40,41^ The trend-level coagulation signal may reflect the reparative arm of the hemostatic system, as low concentrations of thrombin are involved in extracellular matrix remodeling and can activate TGF-β.^42,43^ At the protein network level, a VEGFA/AKT/ERK1/2 hub with downstream suppression of immune surveillance nodes (CD83, IL15RA, CD55) and NOTCH2-mediated neurovascular remodeling corroborated TGF-β-driven resolution.

Time since injury distributions did not differ between ISOVF_high groups (KS p = 0.97), suggesting these differences reflect variability in inflammatory resolution pace rather than recovery phase. If confirmed in longitudinal analyses, the combination of high ISOVF with inflammatory pathway downregulation and early TGF-β signaling could represent a non-invasive marker of favorable recovery trajectory. The lower PPCS rate in ISOVF_high Group 2 (**Results**), though not significant, is directionally consistent with this resolution profile. However, the cross-sectional design precludes causal inference, and larger longitudinal samples are needed to test whether early proteomic evidence of inflammatory resolution paired with elevated free water predicts favorable outcomes.

Across all participants and metrics, the corona radiata, cingulum, superior longitudinal fasciculus, sagittal stratum, and external capsule were most commonly affected, with the corticospinal tract consistently most resilient (**Supplementary Figure 3**). This could reflect less biomechanical vulnerability of projection tracts relative to commissural and association fibers. Direct comparison with prior dMRI-inflammation studies is complicated by biological differences (animal models, chronic disease, varying inflammatory phases), but our data support this broader metric-specific pattern.

The 7- to 35-day window captures a critical transitional period in the post-injury immune response. Experimental models demonstrate that activated microglia exhibit a bimodal response during this timeframe, with an initial reparative M2-like peak around 7 days followed by a pro-inflammatory M1-like peak at 21–28 days,^15^ when inflammation may either resolve toward repair or progress toward maladaptive chronic states. Identifying coordinated pathway activations in the subacute phase may eventually help characterize an adolescent’s inflammatory state non-invasively, informing whether they are on a restorative trajectory or warrant closer monitoring. WM myelination continues into the mid-20s^44^ and complement-mediated synaptic pruning remains active during this period,^45^ so concussion-related complement activation may interact with developmental remodeling. The associations identified here may be specific to adolescence.

Mapping these distinct inflammatory profiles to clinical phenotypes is a critical next step. While emerging evidence links peripheral biomarkers to overall post-concussive symptom severity and to specific somatic, cognitive, or affective domains,^46–51^ directly mapping central neuroinflammation remains severely limited by the impracticality of molecular imaging (e.g., TSPO PET) in children. By demonstrating that non-invasive diffusion MRI metrics are sensitive to distinct inflammatory states, our findings provide a methodological bridge: future clinical trials can match targeted interventions to a patient’s specific biological state, addressing the historical failure of broad anti-inflammatory agents in TBI.

## Strengths and Limitations

A key strength of this study is the use of an unbiased, pathway-level proteomic approach to explore inflammatory processes associated with diffusion MRI metrics, rather than restricting analyses to a small set of candidate biomarkers. The multi-site design increases generalizability but introduces site-related variability, which we addressed through ComBat harmonization and covariate adjustment; residual site effects cannot be entirely excluded. The sample size is modest, and the same participant can fall into different groups across metrics. Individual proteins did not survive FDR correction, but GSEA is designed to detect coordinated pathway-level shifts even when individual protein effects are subtle.

Peripheral blood proteomic profiling reflects systemic rather than brain-specific inflammatory activity, and Olink PEA reports relative protein expression (NPX) rather than absolute concentrations, limiting direct clinical translation. The 7- to 35-day collection window, while clinically relevant, spans a period of rapidly evolving inflammatory processes, and timing variability across participants may influence the observed associations. The study did not account for pubertal stage or menstrual cycle phase, which may influence circulating protein expression and dMRI. Proteomic findings were not validated in wet lab, and IPA networks represent literature-derived interactions rather than causal relationships. Finally, the cross-sectional design precludes conclusions about whether the inflammatory profiles drive the diffusion changes, result from them, or reflect concurrent processes.

## Conclusions

We evidence that dMRI metrics are differentially sensitive to distinct inflammatory states in the subacute period after adolescent concussion. Metrics capturing tissue disorganization were associated with hemostatic-inflammatory upregulation consistent with vascular involvement, metrics reflecting reduced tissue complexity and neurite density with interferon-driven immune activation, and elevated free water with inflammatory resolution signaling. These associations were not explained by demographic differences or time since injury, suggesting genuine biological heterogeneity in the post-injury response. Beyond grounding long-hypothesized links between specific dMRI metrics and inflammatory mechanisms at the pathway level, these findings suggest peripheral proteomic profiling may identify in which phase of the inflammatory cascade a patient is. The failure of broad anti-inflammatory trials in TBI likely reflects this heterogeneity: targeted interventions will require matching treatment to inflammatory phenotype.

## Data Availability

Data may be made available through a request to the authors

## Acknowledgements

This work was conducted on behalf of the Concussion Assessment, Research, and Education for Kids (CARE4Kids) consortium. We thank all members for their contributions and support. The authors would like to thank the adolescents and families who participated in the CARE4Kids study. The authors used AI-assisted tools during preparation of this manuscript. Anthropic’s Claude was used to condense and edit author-written text for length and clarity, to generate the participant flow (CONSORT) diagram from author-provided data (Figure 2), and to assist in writing figure-generation code. It was also used as a brainstorming aid to help understand potential biological mechanisms linking inflammatory pathways to diffusion MRI metrics; all synthesis was drafted and edited by authors with relevant domain expertise. Google’s Gemini was used to produce part of the potholes/peaks figure (Figure 1). All AI-assisted text, figures, and code were reviewed and edited by the authors, who take full responsibility for the accuracy and integrity of the manuscript. The scientific interpretations and conclusions are the authors’ own.

## Supplementary Materials

**Supplementary Table 1.**
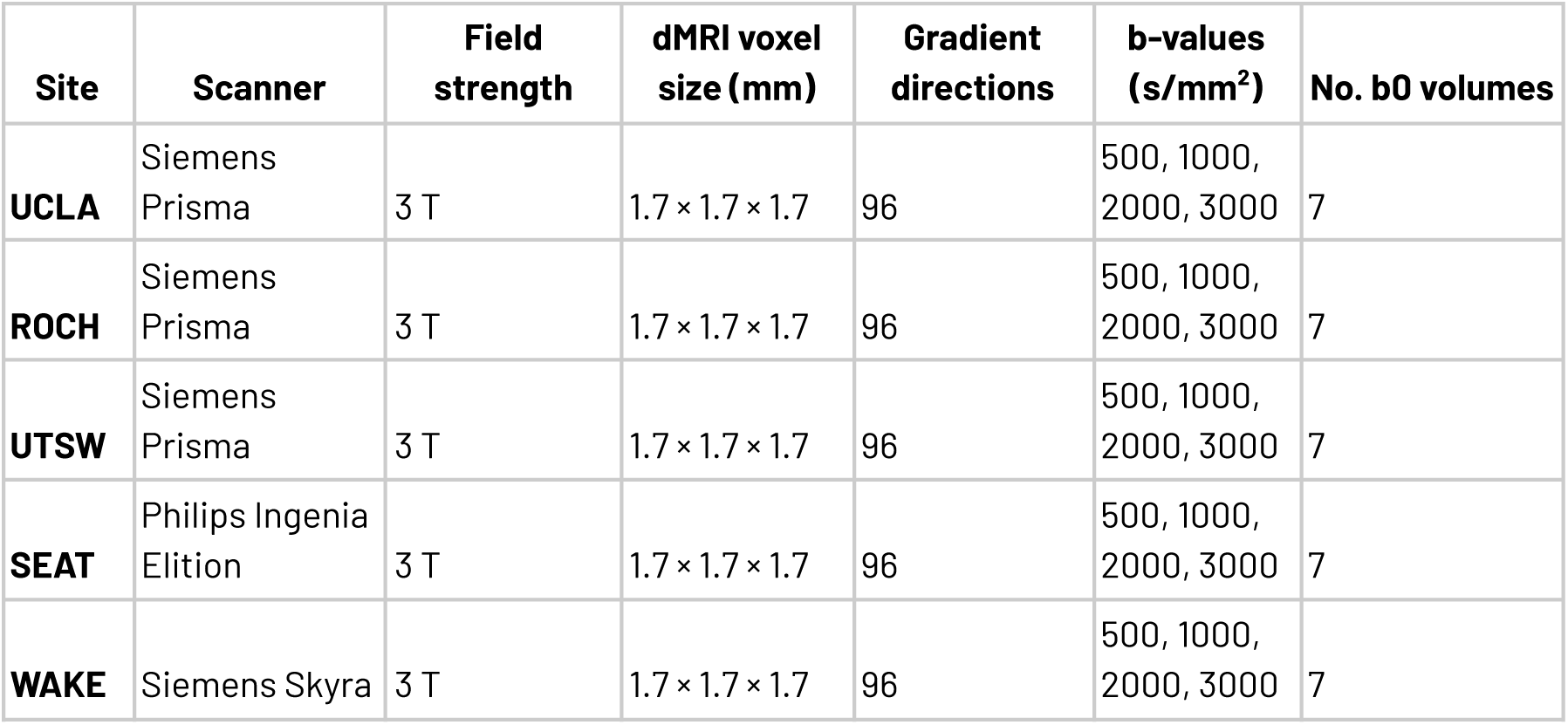
dMRI imaging parameters across sites. For the 5 sites that collected dMRI data the scanner manufacturer and model are shown. Field strength, voxel size,gradient directions, b-values and number of b0 volumes were consistent across sites.

## Supplementary Methods

### MRI Processing

Diffusion-weighted images were processed using a custom pipeline built on FSL (v6.0.5^29^), MRtrix3,^30^ DIPY (v1.11.0^31^), and the dmipy toolbox.^32^ <u>Denoising and artifact correction</u>: Raw diffusion data first underwent denoising using Marchenko-Pastur PCA (MP-PCA) as implemented in MRtrix3’s dwidenoise,^52^ which exploits the redundancy of diffusion-weighted signals across volumes to suppress thermal noise. Gibbs ringing artifacts were then removed using the sub-voxel shift method (mrdegibbs^53^). <u>Susceptibility distortion correction</u>: Susceptibility-induced geometric distortions were estimated from AP-PA fieldmaps and corrected using FSL’s topup^54^ with the standard b02b0.cnf configuration. Spatial dimensions of fieldmap and diffusion volumes were reconciled by trimming to the minimum common matrix size when necessary. <u>Eddy current and motion correction</u>: Eddy current–induced distortions and inter-volume subject motion were corrected using FSL’s eddy_openmp.^55^ Topup field estimates were incorporated into the eddy correction step. A brain mask for eddy was generated using FSL’s Brain Extraction Tool (BET^56^) with a fractional intensity threshold of 0.3. The b-vector orientations were rotated to account for motion correction. <u>Bias field correction</u>: Residual B1 field inhomogeneities were corrected by estimating the bias field from the first b=0 volume using ANTs’ N4 bias field correction^57^ and dividing it out across all volumes.

<u>DTI model fitting</u>: The diffusion tensor was fit to the preprocessed data using FSL’s dtifit,^58^ yielding maps of fractional anisotropy (FA), mean diffusivity (MD), axial diffusivity (AD), and radial diffusivity (RD). <u>DKI</u>: The diffusion kurtosis tensor was fit using DIPY’s DiffusionKurtosisModel,^31,59^ yielding maps of kurtosis FA (KFA), mean kurtosis (MK), axial kurtosis (AK), and radial kurtosis (RK). <u>NODDI</u>: NODDI parameters were estimated using the Watson distribution model as implemented in the dmipy framework.^21,32^ Input data were thresholded to remove negative values prior to fitting. The model produced maps of the intracellular volume fraction (ICVF), orientation dispersion index (ODI), and isotropic volume fraction (ISOVF).

### Proteomic Data Processing

Plasma samples were aliquoted into a pre-assigned well on a plate, then transferred to a sample source plate and diluted. The immune reaction or incubation was performed next, during which high-multiplex PEA™ probes, matched pairs of antibodies with unique DNA oligonucleotides, bind to their respective proteins in the samples. This incubation period was 18 hours for this study. Following the incubation, oligonucleotides brought into proximity hybridized and were extended. The DNA barcode was then amplified by Polymerase Chain Reaction (PCR). Unique sample indexes were added to each sample to enable pooling of DNA amplicons across all samples. The final DNA amplicons in the Olink Explore HT libraries included specific barcode sequences for each assay, sample-specific indexes, and required sequences for Illumina sequencing. Following the PCR procedure, PCR pooling was performed, which combines samples from the same dilution block into a single pool, yielding 1 pool per dilution block, each containing 192 samples/controls. Library purification and quality control were performed using AMPure® XP Magnetic Beads to purify the samples and the Agilent® Tapestation 4200 instrumentation and High Sensitivity D1000 kits to assess sample quality. Following library purification, samples were pooled and sequenced using next-generation sequencing on the Illumina NovaSeq X Plus platform with single-end 24-base reads, in accordance with the Olink Explore HT protocol. Relative concentrations of each biomarker, based on matched counts (the number of reads for each specific combination of sample and assay), were calculated using the Olink Explore HT software. The measured raw protein expression levels were normalized using the intensity normalization method to yield Normalized Protein eXpression (NPX) values on a log2 scale. Standard quality control procedures were applied according to manufacturer recommendations.

### Ingenuity Pathway Analysis

Protein networks were generated using Ingenuity Pathway Analysis (IPA) software from a subset of MRI metrics that exhibited distinct pathway profiles. Proteins that passed the unadjusted p-value cutoff (<0.05) were uploaded to IPA software for core analysis (QIAGEN Inc., https://digitalinsights.qiagen.com/IPA) for exploratory purposes. These data-driven protein networks were displayed with nodes and lines based on references from the literature, textbooks, or canonical data in the human gene knowledge base. The color and intensity of the nodes indicate the level of dysregulation (red for upregulation, green for downregulation) and predicted activation status (orange for activated, blue for inhibited). The relationships between nodes are represented by solid lines (direct) or dashed lines (indirect).

**Supplementary Figure 1.**
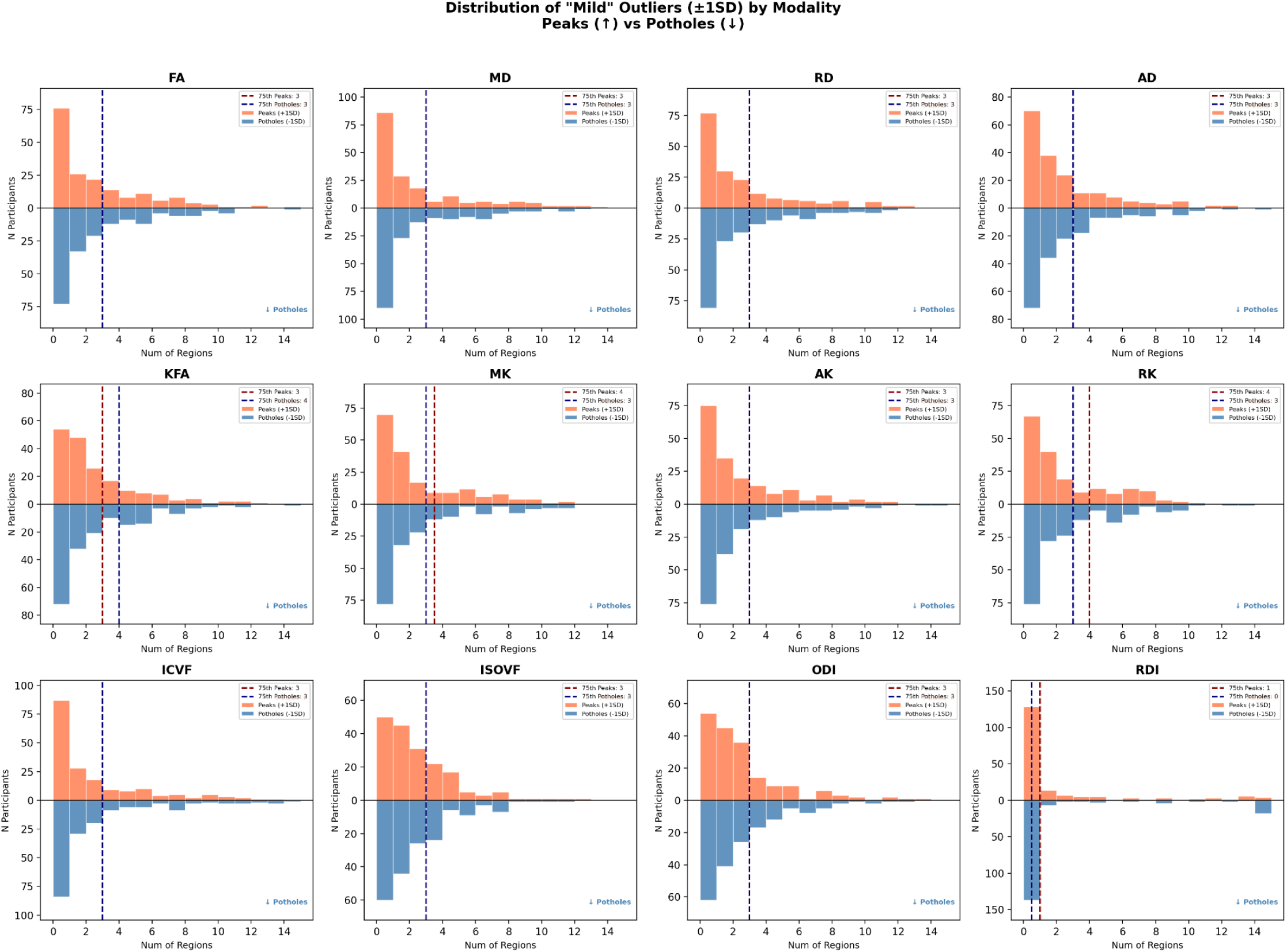
Histograms of potholes and peaks across metrics. Histograms are shown for all 12 metrics, with the number of regions with Z-scores above 1 (peaks) on top in coral and the number of regions with Z-scores below −1 (potholes) mirrored below in blue.

**Supplementary Figure 2.**
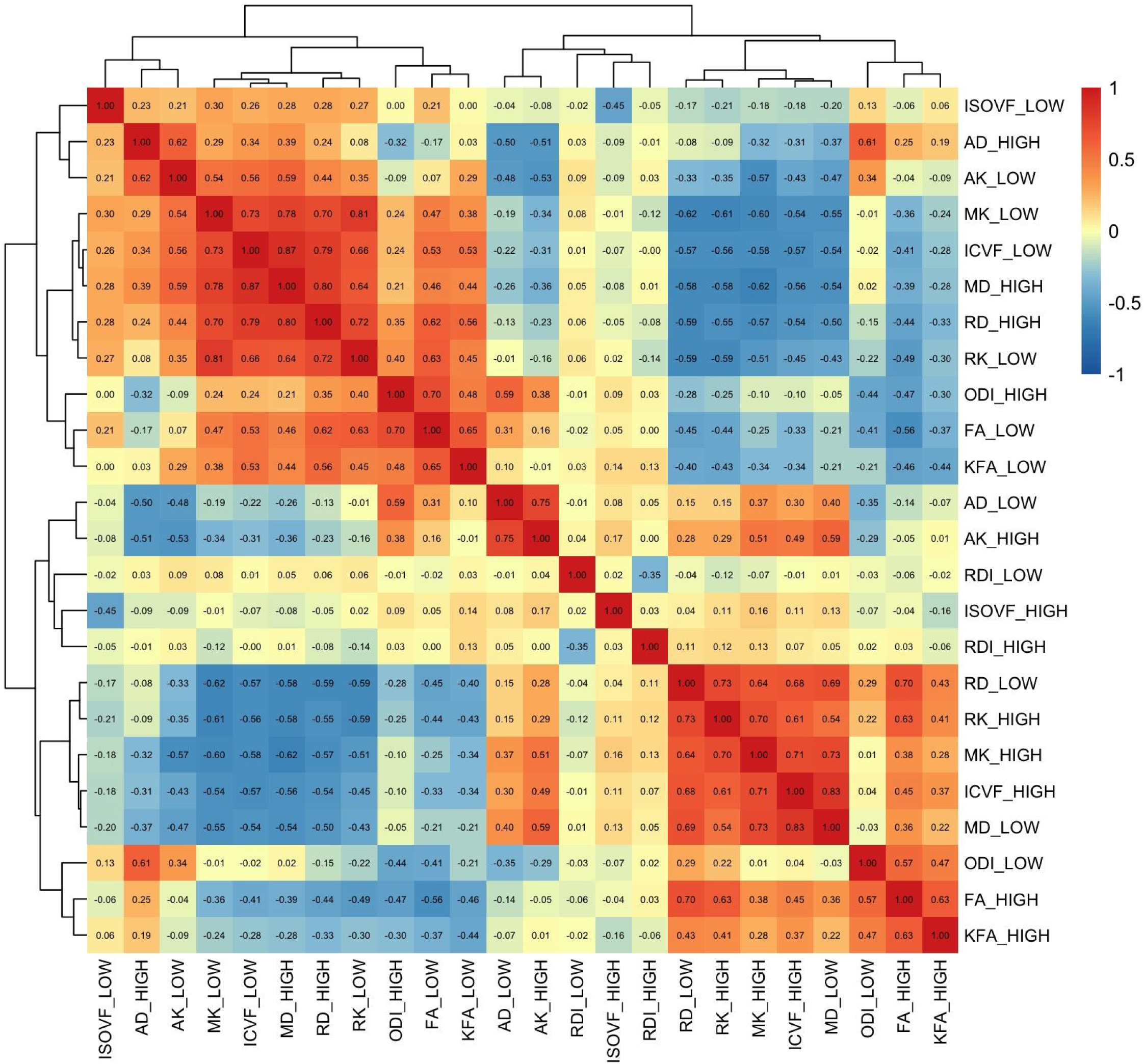
Correlations among dMRI groupings. Pairwise Pearson correlations are shown for all 24 groupings (12 metrics × high and low z-score groups) with correlation coefficients displayed in each cell and color-coded according to the scale bar (red = positive, blue = negative). Groupings are clustered hierarchically based on their pairwise correlation structure, revealing two primary clusters: one comprising diffusivity-based metrics in their expected directions (AD_HIGH, AK_LOW, MK_LOW, ICVF_LOW, MD_HIGH, RD_HIGH, RK_LOW, ODI_HIGH, FA_LOW, KFA_LOW), and a second comprising their opposing groupings (RD_LOW, RK_HIGH, MK_HIGH, ICVF_HIGH, MD_LOW, FA_HIGH, KFA_HIGH), with DSI-derived metrics (RDI, ISOVF) and ODI_LOW showing weaker correlations with both clusters. Metrics: AD, axial diffusivity; AK, axial kurtosis; FA, fractional anisotropy; ICVF, intracellular volume fraction; ISOVF, isotropic volume fraction; KFA, kurtosis fractional anisotropy; MD, mean diffusivity; MK, mean kurtosis; ODI, orientation dispersion index; RD, radial diffusivity; RDI, restricted diffusion imaging; RK, radial kurtosis.

**Supplementary Figure 3.**
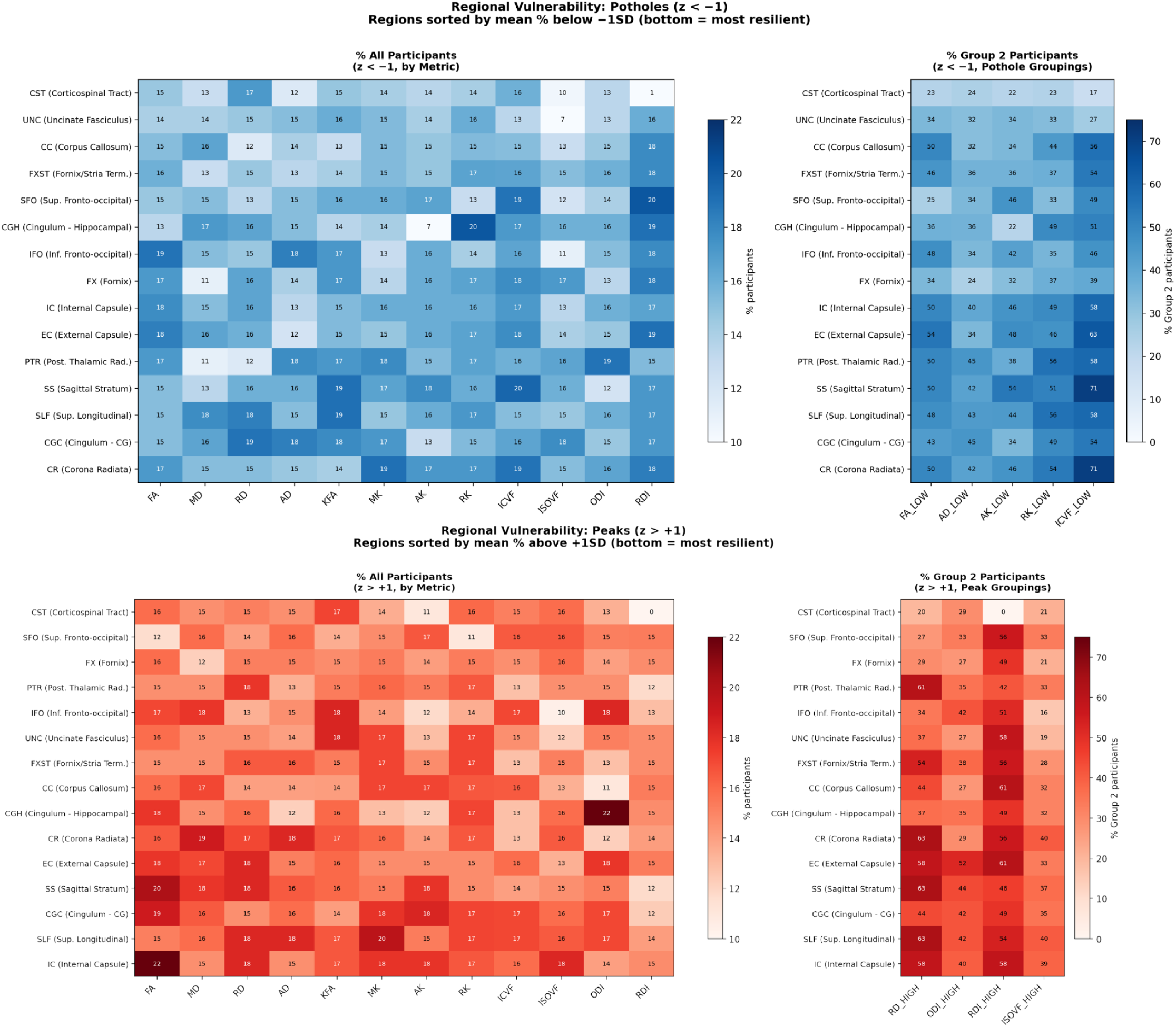
Regional distribution of potholes and peaks. Heatmaps showing the percentage of participants with outlier values in each of 15 WM ROIs across 12 diffusion metrics, separated by direction. Upper panels show potholes (z < −1); lower panels show peaks (z > +1). Left panels in each row show the percentage of all participants with outlier values in the given direction for each metric × region combination, with regions sorted from most resilient (bottom) to most vulnerable (top) based on mean percentage across metrics. Right panels show the percentage of Group 2 participants — those with the most widespread outlier values — with outlier values in the expected direction for each of the primary imaging groupings (pothole groupings: FA_LOW, AD_LOW, AK_LOW, RK_LOW, ICVF_LOW; peak groupings: RD_HIGH, ODI_HIGH, RDI_HIGH, ISOVF_HIGH). Color intensity reflects the percentage of participants with outlier values in the relevant direction, ranging from 10–22% in the all-participants panels and 0–75% in the Group 2 panels. Regions: CC = corpus callosum; CGC = cingulum (cingulate gyrus); CGH = cingulum (hippocampal); CR = corona radiata; CST = corticospinal tract; EC = external capsule; FX = fornix; FXST = fornix/stria terminalis; IC = internal capsule; IFO = inferior fronto-occipital fasciculus; PTR = posterior thalamic radiation; SFO = superior fronto-occipital fasciculus; SLF = superior longitudinal fasciculus; SS = sagittal stratum; UNC = uncinate fasciculus.

**Supplementary Figure 4.**
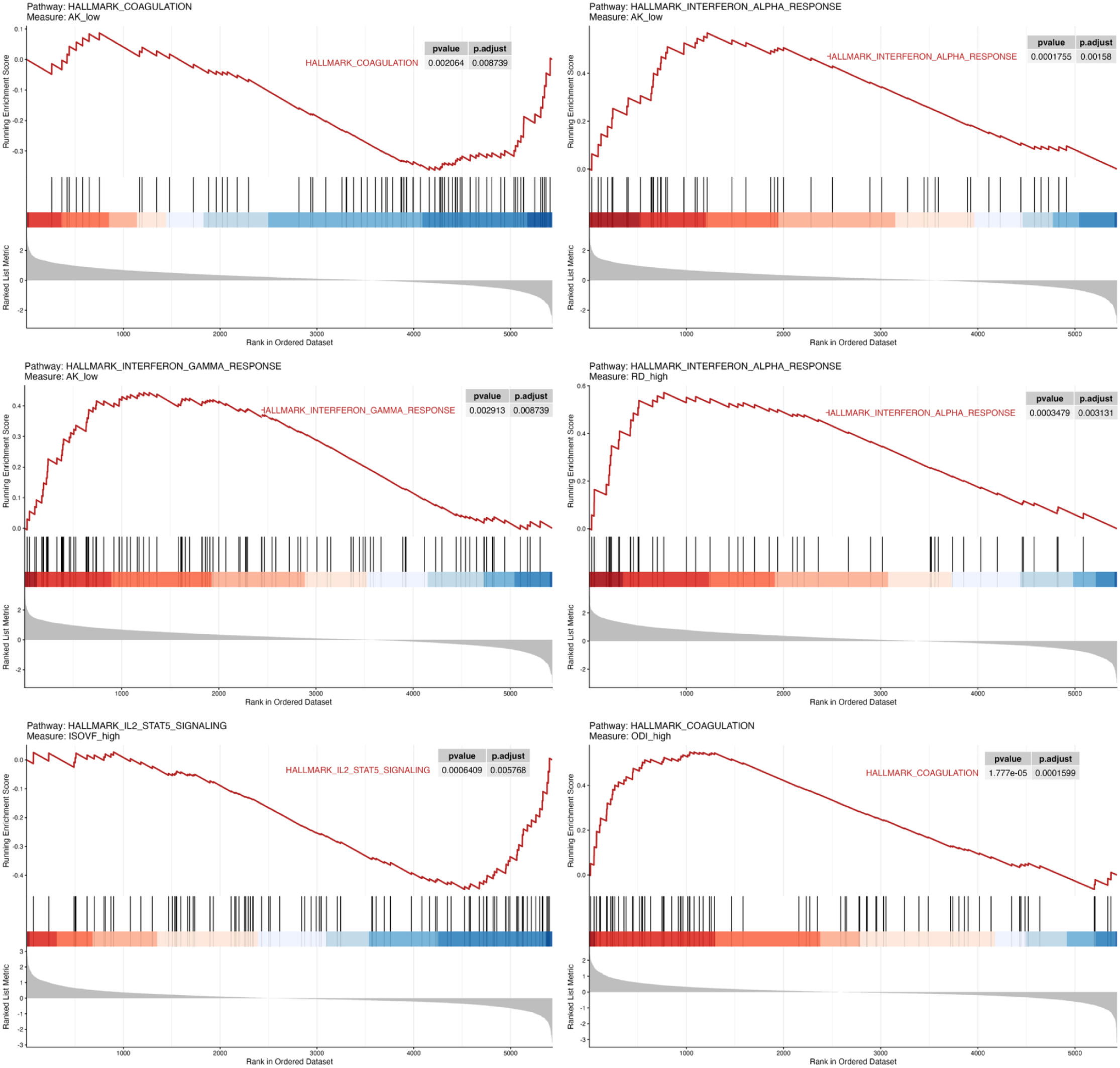
Enrichment plots for significant pathways (q<0.01)

